# Screening of SARS-CoV-2 among homeless people, asylum-seekers and other people living in precarious conditions in Marseille, France, March–April 2020

**DOI:** 10.1101/2020.05.05.20091934

**Authors:** Tran Duc Anh Ly, Van Thuan Hoang, Ndiaw Goumballa, Meriem Louni, Naomie Canard, Thi Loi Dao, Hacene Medkour, Audrey Borg, Kevin Bardy, Véra Esteves-Vieira, Véronique Filosa, Bernard Davoust, Oleg Mediannikov, Pierre-Edouard Fournier, Didier Raoult, Philippe Gautret

## Abstract

Surveillance of SARS-CoV-2 infection among sheltered homeless and other vulnerable people might provide the information needed to prevent its spread within accommodation centres. In March-April, we enrolled 411 homeless individuals, 77 asylum-seekers, 58 people living in precarious conditions, and 152 employees working in these accommodation centres and collected nasal samples. SARS-CoV-2 carriage was assessed by quantitative PCR. Overall, 49 (7.0%) people were positive for SARS-CoV-2, including 37 homeless individuals (of 411, 9.0%), 12 employees (of 152, 7.9%). SARS-CoV-2 positivity correlated with symptoms, although 51% of positive patients did not report respiratory symptoms or fever. Among homeless people, being young (18-34 years) (OR: 3.83 [1.47-10.0], p=0.006) and being housed in one specific shelter (OR: 9.13 [4.09-20.37], p<0.0001) were independent factors associated with the SARS-CoV-2 positivity rates (11.4% and 20.6%, respectively). The survey reveals the role of collective housing in relation to viral transmission within centres.

## Introduction

Since March 2020, the coronavirus disease (COVID-19) caused by severe acute respiratory syndrome coronavirus 2 (SARS-CoV-2) has spread over more than 200 countries and territories worldwide *(1)*. Homeless people are a vulnerable group who may potentially be exposed to this infection and have potentially more severe outcomes than in the general population, due to their poor living conditions, the higher prevalence of comorbidity, and mental and physical conditions impaired by substance or alcohol abuse *(2-6)*. Crowded conditions in shelters without specific preventive measures could facilitate viral transmission *(7, 8)*. In several U.S. cities, 1,192 residents and 313 staff members were tested in 19 homeless shelters in March–April and high rates of SARS-CoV-2 carriage were observed in residents (25%) and staff members (11%) *(9, 10)*; the prevalence was also reported to be 9.7-15.5% and 13.3%-14.8% among residents and staff members in within 3 homeless shelters in Washington, respectively *(11)*. This raised concerns that the virus may be widely transmitted within homeless shelters, even when infection control vigilance is high.

Over the past two decades, our institute has carried out a large number of surveys among homeless persons within two shelters (A and B) in Marseille, France. We observed a high prevalence of respiratory symptoms and signs *(12)* and high carriage rates of both respiratory viruses *(13)* and bacteria *(14)*. suggesting that SARS-CoV-2 infection might also be frequent in this population. Based on the preliminary information that some homeless persons from these two shelters presented with COVID-19 symptoms, we organised a screening campaign in collaboration with the staff in charge of these shelters. We subsequently received other requests for screening from several accommodation centres specialising in housing vulnerable people. In this study, we present the results of SARS-CoV-2 screening campaigns conducted among sheltered homeless individuals, in comparison with asylum-seekers, other persons living in precarious conditions, and employees working in the accommodation centres. We also investigated the role of potential risk factors for virus carriage among the homeless population.

## Methods and materials

### Ethics

Ethical approval was obtained from the Institutional Review Board and Ethics Committee of our institute (2020-015).

## Setting, study design and population

A cross-sectional survey was conducted between 26 March and 17 April 2020 in different populations including homeless people residing in four shelters (A-D) and four hotels (1-4), other people living in precarious condition (housed in residences α and β), asylum-seekers (housed in residence γ), and employees working in these accommodation centres.

Homeless shelters (A-C) include emergency (overnight stay) units with a rapid turnover (7–14 nights), and special (permanent stay) units dedicated to high-risk sedentary homeless persons characterised by a high level of poverty, poor hygiene, alcoholism, mental illness and chronic diseases. Shelters A and B are for men only while shelter C is for women only. Shelter D houses male and female homeless people and offers the possibility to keep their pets when needed. Characteristics of the facilities are described in Table 1. All residents of homeless shelters were placed under strict lockdown since 17 March, in line with the whole French population (=C0), allowing all homeless people to stay in the shelter 24 hours a day. The male population of Shelter B (initial group B) was sub-divided into three groups by the staff of the facility, in order to avoid overcrowding: i) elderly people, those with reduced mobility and those needing medical care were kept in Shelter B, ii) people aged 18-45 were progressively moved to Hotel 1 from C0 to C7, iii) people aged 30-80 years were moved to Hotel 2 from C7 to C14. Similarly, the female population of Shelter C (initial group C) was sub-divided into three groups: elderly people, those with reduced mobility and those needing medical care were kept in Shelter C, ii) pregnant women and those with mental illness were moved to Hotel 3 at C0; iii) others were moved to Hotel 4 at C0. All residents moved to hotels have been kept under relatively strict lockdown since C0, with the exception of the day of transfer.

**Table 1.** Characteristics of shelters, hotels and residences

| Descriptive | Homeless shelters and hotels |  |  |  |  |  |  |  | Residence for specific populations living in precarious conditions |  | Residence for asylum-seekers |
| --- | --- | --- | --- | --- | --- | --- | --- | --- | --- | --- | --- |
|  |  |  |  |  |  |  |  |  | Residence α | Residence β | Residence γ |
|  | Shelter A | Shelter B | Shelter C | Shelter D | Hotel 1 | Hotel 2 | Hotel 3 | Hotel 4 |  |  |  |
| Type of residents | Adults, males | Adults, males | Adults, females | Adults, males/ females and pets | Adult males | Adult males | Adult females | Adult females | Adults, males/females (drug addiction, chronic diseases...) | Teenage mothers and their children | Family groups<sup>1</sup> |
| Total capacity/emergency beds/long-term beds | 283/ 248/ 35 | 310/ 260-280/ 30-50 | 64/ 50/14 | 33/ NA/NA | 70/NA/NA | 100/NA/NA | 15/NA/NA | 10/NA/NA | 20/NA/NA | 20/NA/NA | 50/NA/NA |
| Room or apartment | 2-3 people/ room | 3-8 people/ room | 2-12 people/ room | 1 person/ room | 2-3 people/ room | 2-3 people/ room | single room | single room | 1-2 people/ apartment | 1 mother-child(ren) pair/ apartment | Family apartments |
| Bathroom and toilets | Collective | Collective | Collective | Private | Collective | Private | Private | Collective | Private | Private | Private |
| Kitchen | Collective | Collective | Collective | Collective | Collective | Collective | Collective | Collective | Private | Private | Private |
| Open space | Large terrace, cultural hall | Large terrace | Large terrace | Large terrace, cultural hall | Large terrace | Large terrace | None | Large terrace | None | Cultural hall, garden | None |
| Lockdown Time between first day of lockdown and screening (days) | 9 | 14 | 15 | 31 | 17 | 20 | 14 | 14 | 24 | 28 | 17 |

Abbreviation: NA not applicable.
<sup>1</sup> single individuals were housed on the basis of two individuals/apartment

Residence α is dedicated to individuals characterised by a high level of poverty, poor hygiene, alcoholism, mental illness and chronic diseases including drug addiction. Residence β specialises in housing teenage mothers and their children. Residence γ is dedicated to asylum-seekers, including family groups and single individuals. All three residences offer long-time housing and all residents have been kept under strict lockdown since C0.

Employees of the different facilities working in different sectors (management staff, social workers, nurses, cleaning staff, catering staff and security staff) returned to their homes on a daily basis after finishing work.

### Screening for Covid-19

Participants were encouraged by the management staff of the facilities to get tested and were then recruited on a voluntary basis. They were systematically asked to provide basic demographic information (sex, age, country of origin), chronic conditions, and any respiratory symptoms or fever in the two weeks prior to sampling. Body temperature was measured using a forehead infrared thermometer. Nasal samples were systematically collected on transport media using Sigma Transwabs (Medical Wire, Corsham, United Kingdom). For self-sampling, participants were invited to insert the swab into their nostrils (about 2 cm). If individuals were unable to perform self-sampling, trained investigators carried out the sampling. Specimens were immediately processed for SARS-CoV-2 PCR testing. Homeless peoples’ pets were also tested with the approval of their owner and their nasal swabs were collected by vets.

### PCR assay

Real-time reverse transcription-PCR amplification was used to confirm the presence of SARS-CoV-2 RNA targeting the gene coding for the envelope (E) protein, as previously described *(15)*. Results were considered positive when the cycle threshold (CT) value of real-time PCR was ≤35.

### Statistical analysis

Statistical procedures were performed using STATA 11.1 software (StataCorp LLC, USA). Percentage differences were tested using Pearson’s chi-square or Fisher’s exact tests when appropriate. Means of quantitative data were compared using Student’s t-test. A p value <0.05 was considered statistically significant. A separate multivariate logistical regression analysis was used to identify independent risk factors for SARS-CoV-2 carriage prevalence among all individuals and in selected groups (when positive cases were found). The results were presented by percentages and odd ratio (OR) with 95% confidence interval (95%CI). The initial model included variables presenting a p-value <0.2. The stepwise regression procedure and likelihood-ratio tests were applied to determine the final model.

## Results

### Participant characteristics (Tables 1, 2 and 3)

Overall, 885 individuals were present in the various facilities at the time of enrolment, including 716 residents and 169 employees (Table 2). A total of 698 (78.9%) subjects agreed to be tested, including 411/698 homeless people (58.9%), 58 non-homeless people living in precarious conditions (8.3%), 77 asylum-seekers (11.0%), and 152 employees (21.8%). Overall, 38.7% were enrolled before C14, 45.9% between C14 and C20, and 15.4% at C21 and later (Table 1, 3). The overall acceptation rate of SARS-CoV-2 testing varied significantly according to the housing facility, ranging from 41.7 to 91.7%. The overall acceptation rate among homeless individuals was 74.6% and was significantly lower than that of employees working in the homeless centres (88.7%, p=0.0008). The acceptance rate among people housed in other facilities, varied from 75.5 to 100% and tented to be lower than that of employees in these facilities.

**Table 2.** Number of screened individuals and results of SARS-CoV-2 PCR detection, according to housing facility

| Housing structures | Number of people present at the time of enrolment |  |  | Number of tested people (%)<sup>1</sup> |  |  |  | Numbers of people testing positive for SARS-CoV-2 (% of tested people) |  |  |  |
| --- | --- | --- | --- | --- | --- | --- | --- | --- | --- | --- | --- |
|  | Total | Residents | Employees | Total | Residents | Employees | p-value | Total | Residents | Employees | p-value |
| **Total** | **885** | **716** | **169** | **698 (78.9)** | **546 (76.2)** | **152 (90.0)** | **0.0001** | **49 (7.0)** | **37 (6.8)** | **12 (7.9)** | **0.77** |
| **Homeless shelters** | **683** | **551** | **132** | **528 (77.3)** | **411 (74.6)** | **117 (88.7)** | **0.0008** | **48 (9.1)** | **37 (9.0)** | **11 (9.4)** | **1.00** |
| Shelter A | 305 | 262 | 43 | 270 (88.5) | 227 (86.6) | 43 (100) | 0.007 | 15 (5.6) | 9 (4.0) | 6 (14.0) | 0.02 |
| Shelter B | 121 | 86 | 35 | 78 (64.5) | 54 (62.8) | 24 (68.6) | 0.7 | 13 (16.7) | 10 (18.5) | 3 (12.5) | 0.7 |
| Shelter C | 48 | 19 | 29 | 40 (83.3) | 14 (73.7) | 26 (89.7) | 0.23 | 2 (5.0) | 0 | 2 (7.7) | NA |
| Shelter D<sup>2</sup> | 36 | 27 | 9 | 33 (91.7) | 25 (92.6) | 8 (88.8) | 1.00 | 0 | 0 | 0 | NA |
| Hotel 1 | 72 | 65 | 7 | 30 (41.7) | 23 (35.4) | 7 (100) | 0.001 | 9 (30.0) | 9 (39.1) | 0 | 0.07 |
| Hotel 2 | 75 | 70 | 5 | 54 (72) | 49 (70) | 5 (100) | 0.3 | 7 (13.0) | 7 (14.3) | 0 | 0.48 |
| Hotel 3 | 14 | 13 | 1 | 12 (85.7) | 11 (84.6) | 1 (100) | 1.00 | 1 (8.3) | 1 (9.1) | 0 | NA |
| Hotel 4 | 12 | 9 | 3 | 11 (91.7) | 8 (88.9) | 3 (100) | 1.00 | 1 (9.1) | 1 (12.5) | 0 | NA |
| **Residence for specific populations living in precarious conditions** | **81** | **63** | **18** | **75 (92.6)** | **58 (92.1)** | **17 (94.4)** | **1.00** | **0** | **0** | **0** | **N/A** |
| Residence $\alpha$ | 28 | 23 | 5 | 28 (100) | 23 (100) | 5 (100) | 1.00 | 0 | 0 | 0 | N/A |
| Residence $\beta$ | 53 | 40 | 13 | 47 (88.6) | 35 (87.5) | 12 (92.3) | 1.00 | 0 | 0 | 0 | N/A |
| **Residence for asylum-seekers** | **121** | **102** | **19** | **95 (78.5)** | **77 (75.5)** | **18 (94.7)** | **0.07** | **1 (1.1)** | **0** | **1 (5.6)** | **N/A** |
| Residence $\gamma$ | | | | | | | | | | | |

Abbreviation: NA : Not applicable;
<sup>1</sup> Acceptance rate<sup>2</sup> Two dogs belonging to two different homeless people in Shelter D were tested and were negative.
Grey cells: four groups in study : Homeless people (N=411); other specific population living in precarious conditions (N=58), asylum seekers (N=77), and employees (N=152) and SARS-CoV-2 prevalence in each group.

**Table 3.**
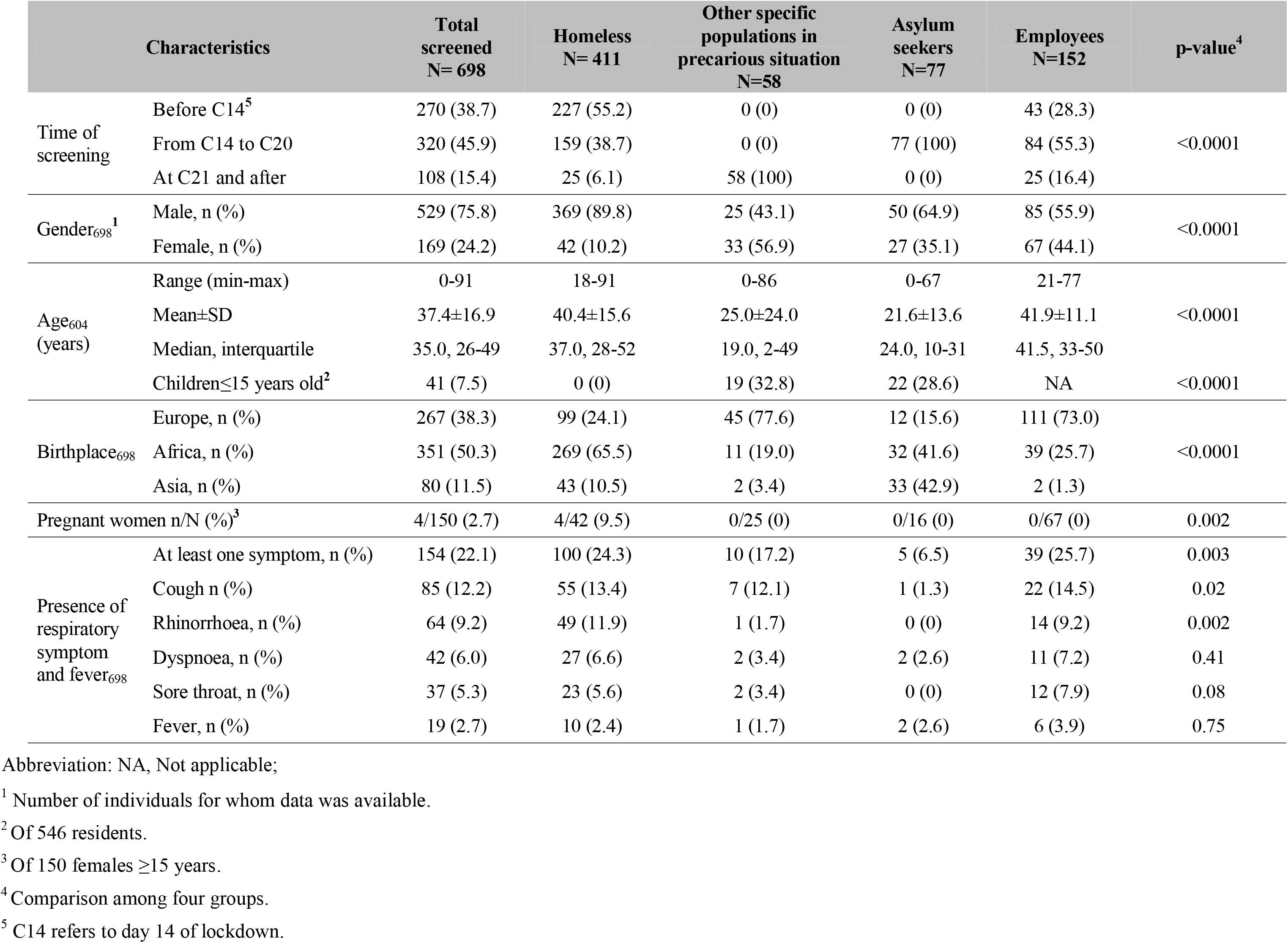
Characteristics of different populations studied

The socio-demographic characteristics of the different populations are presented in Table 3. The male to female gender ratio was 3:1 and the median age was 35.0 years (ranging from 0 to 91 years) with significant variations among different populations. A male predominance was observed among homeless persons and asylum seekers. Children ≤15 years old accounted for 7.5% of all residents. Two-thirds of individuals were migrants. A predominance of African origin was found among homeless individuals, while other people living in precarious conditions and employees were more likely to be European. There were only four pregnant women (between 26 and 36 weeks of pregnancy), all housed in Hotel 3.

Regarding clinical findings, among all the participants, 22.1% reported at least one respiratory symptom or fever with significant variations among different populations. The highest prevalence was observed among employees (25.7%) and homeless persons (24.3%). A cough was the most commonly reported symptom (32.7%) followed by rhinorrhoea (20.4%), dyspnoea (12.2%) and fever (12.2%). No deaths were reported during the study period.

### SARS-CoV-2 detection (Table 2, 4, 5)

In total, 49 participants (7.0%) tested positive for SARS-CoV-2, including 37 homeless people (of 441, 9.0%) and 12 employees (of 152, 7.9%, including seven security staff from Shelters A, B and C and residence γ, four nurses from Shelter A and one management staff member from Shelter C). Only two female homeless people tested positive, including one woman who was 36 weeks pregnant and who frequently attended the hospital during the lockdown and one person with mental illness who did not comply with lockdown measures.

Two dogs belonging to two different homeless people in Shelter D tested negative. With regard to the housing facilities, the highest SARS-CoV-2 positivity rates were observed in homeless persons in Hotel 1 (39.1%), in Shelter B (18.5%) and in Hotel 2 (14.3%). Among employees, the highest positive rates were in those working at homeless Shelters A (14%) and B (12.5%).

Of the 49 SARS-CoV-2-positive participants, 51.0% were asymptomatic. Positive participants were more likely to be symptomatic compared to negative participants (Odd-ratio OR=3.8 95%CI [2.1-6.9], p<10^-4^). There was no significant difference of PCR CT values between asymptomatic (mean CT [±SD]: 26.9±5.0) and symptomatic individuals (25.7±5.4, p=0.43). The overall proportion of asymptomatic carriers among all tested individuals was 3.6% and that of symptomatic carriers was 3.4%.

Table 3 shows SARS-CoV-2 positivity rates among homeless people according to the time of screening, demographics and housing facility, using univariate analysis. No significant differences were observed according to gender and country of origin regarding SARS-CoV-2 positivity rates. Screening between C14 and C20 and screening in the group B population (Shelter B and hotels to which people from Shelter B were moved) resulted in a significantly higher proportion of positive PCR as compared to screening before C14 or screening in other homeless facilities, respectively. In addition, being young (18-34 years) was associated with an increased prevalence of virus detection. Cough, rhinorrhoea and fever were associated with viral carriage. Using multivariate analysis (Table 4), being young and screening conducted in the group B population remained significantly associated with a higher likelihood of SARS-CoV-2 detection.

**Table 4.** Associations between multiple factors and SARS-CoV-2 positivity among 411 homeless people (univariate and multivariate analysis

| Characteristics |  | Positive<br>N=37 | Negative<br>N=374 | Univariate |  | Multivariate |  |
| --- | --- | --- | --- | --- | --- | --- | --- |
|  |  |  |  | OR [95%CI] | p-value | aOR [95%CI] | p-value |
| Time of screening | Before C14<sup>2</sup> | 9 (4.0) | 218 (96.0) | Ref |  | - | - |
|  | From C14 to C20 | **28 (17.6)** | **131 (82.4)** | **5.17 [2.36-11.31]** | **<10<sup>-4</sup>** | - | - |
|  | At C21 and later | 0 (0) | 25 (100) | NA | 0.6 | - | - |
| Gender | Male, n (%) | 35 (9.5) | 334 (90.5) | Ref |  |  |  |
|  | Female, n (%) | 2 (4.7) | 40 (95.2) | 1.56 [0.48–9.04] | 0.32 |  |  |
| Age (years) | ≥50 | 7 (6.2) | 105 (93.8) | Ref |  | Ref |  |
|  | 35–49 | 9 (8.3) | 100 (91.7) | 1.34 [0.48-3.76] | 0.56 | - | - |
|  | 18–34 | **20 (11.4)** | **156 (88.6)** | **1.92 [0.78-4.7]** | **0.15** | 3.83 [1.47-10.0] | 0.006 |
| Birthplace | Europe, n (%) | 6 (6.0) | 93 (94.0) | Ref |  |  |  |
|  | Africa, n (%) | 27 (10.0) | 242 (90.0) | 1.72 [0.69-4.32] | 0.24 |  |  |
|  | Asia, n (%) | 4 (9.3) | 39 (90.7) | 1.58 [0.42-5.94] | 0.49 |  |  |
| Housing facility<sup>1</sup> | Other homeless facilities | 11 (3.9) | 274 (96.1) | Ref |  | Ref |  |
|  | Group B | **26 (20.6)** | **100 (79.4)** | **6.47 [3.1-13.6]** | **<10<sup>-4</sup>** | 9.13 [4.09-20.37] | <10<sup>-4</sup> |

Abbreviation: Ref, Reference; NA, Not applicable; OR, Odd-ratio; aOR, adjusted Odd-ratio.
<sup>1</sup> Group B includes Shelter B, Hotels 1 and 2; Other homeless facilities include Shelters A, C, D and Hotels 3, 4.
<sup>2</sup> C14 means day 14 of lockdown.
Bold lines indicate the variables recruited in initial multivariate mode.

## Discussion

To our knowledge, this is the only study addressing SARS-CoV-2 carriage among different precarious populations including homeless adults but also children and other hard-to-reach populations during the COVID-19 outbreak in France. The strength of our study is its large population size, with a high (78.9%) acceptance rate toward testing, particularly among individuals living in precarious conditions (92.1%) suggesting that this population is concerned about the disease.

We found an overall 7.0% SARS-CoV-2 positivity rate, with most infected individuals among homeless people and employees working in homeless facilities, while no cases were found in asylum-seekers and in other people also living in precarious conditions. Detection of SARS-CoV-2 correlated with symptoms although many patients who tested positive did not report any respiratory symptoms or fever. Homeless people and professionals in contact with homeless people are therefore at a high risk of COVID-19. These populations should benefit from screening campaigns and specific measures aiming at mitigating the risks of transmission of the disease within these populations and to the overall population should be implemented.

Among the population of the four homeless shelters (A–D) that were screened, the highest prevalence was observed in populations initially housed at Shelter B. This may have resulted from the higher number of individuals per room at this shelter, as compared to other shelters, which may have encouraged transmission of the virus. Sleeping in shared dormitories and using shared bathrooms, toilets and kitchens make the implementation of social distancing measures in the context of homeless shelters particularly challenging. Being young (18-34 years) was an independent factor associated with SARS-CoV-2 detection in the homeless group, which may be due to a higher propensity of younger homeless people to develop social interactions within the shelters and hotels as compared to older people aged ≥50 years.

This work has some limitations. Our study population was not randomly and homogenously recruited. Participants’ medical histories and use of individual preventive measures were not documented. Individuals were not asked about anosmia and ageusia. Notwithstanding these limitations, our data provide a novel insight into the epidemiology of SARS-CoV-2 among different vulnerable urban populations. The survey also reveals the role of collective housing in relation to viral transmission within accommodation centres. Further genomic investigations are needed to better assess the source(s) and mode(s) of transmission of COVID-19 in this context.

## Data Availability

No data availability

## Acknowledgements

This work was supported by the French Government under the “Investments for the Future” programme managed by the National Agency for Research (ANR), Méditerranée-Infection 10-IAHU-03, and was also supported by Région Provence-Alpes-Côte d’Azur. This work had received financial support from the Fondation Méditerranée Infection.

## Conflicts of interest

No potential conflict of interest relevant to this letter was reported.

## Notes

### Competing Interest Statement

The authors have declared no competing interest.

